# Measuring Community Resilience During the COVID-19 based on Community Wellbeing and Resource Distribution

**DOI:** 10.1101/2022.05.23.22275454

**Authors:** Jaber Valinejad, Zhen Guo, Jin-Hee Cho, Ing-Ray Chen

## Abstract

The COVID-19 pandemic has severely harmed every aspect of our daily lives, resulting in a slew of social problems. Therefore, it is critical to accurately assess the current state of community functionality and resilience under this pandemic for successful recovery. To this end, various types of social sensing tools, such as tweeting and publicly released news, have been employed to understand individuals’ and communities’ thoughts, behaviors, and attitudes during the COVID-19 pandemic. However, some portions of the released news are fake and can easily mislead the community to respond improperly to disasters like COVID-19. This paper aims to assess the correlation between various news and tweets collected during the COVID-19 pandemic on community functionality and resilience. We use fact-checking organizations to classify news as real, mixed, or fake, and machine learning algorithms to classify tweets as real or fake to measure and compare community resilience (CR). Based on the news articles and tweets collected, we quantify CR based on two key factors, *community wellbeing* and *resource distribution*, where resource distribution is assessed by the level of *economic resilience, and community capital*. Based on the estimates of these two factors, we quantify CR from both news articles and tweets and analyze the extent to which CR measured from the news articles can reflect the actual state of CR measured from tweets. To improve the operationalization and sociological significance of this work, we use dimension reduction techniques to integrate the dimensions.

## Introduction

### Motivation

The recent outbreak of COVID-19 has disrupted every aspect of our daily lives. To absorb and adapt against COVID-19 in an agile manner and quickly recover from it, maintaining a healthy, socially connected, and prepared community is critical [1]. Community wellbeing is an essential asset to build a resilient community [2]. In addition, how resources are distributed in a community can present the community’s resilience against a disaster like COVID-19. High accessibility to resources and their fair distribution are the keys to community resilience [3, 4]. Numerous sensing tools are available to assess community resilience, including online websites, social media, surveys, and infrastructure sensing. Among these sensing tools, while social media is an essential social sensing tool for revealing community behavior and thought, it has received little attention previously. Seven out of ten Americans use social media to exchange personal information, interact with content, and connect with others [5]. According to a recent research [6], the psychological states of a whole population can be revealed through social media. Social media provides a platform for billions of users to communicate, express sentiments, and provide real-time updates about human interaction on a large scale [7]. Twitter is one of the major community social media platforms. In this regard, numerous studies have employed tweeter to evaluate population behavior [6, 7, 8, 9, 10, 11]. Unfortunately, fake news may negatively impact maintaining community wellbeing and equitable resource distribution during COVID-19. The Internet, social media, and mass media platforms have generated a large volume of information flow during the COVID-19. Part of the information volume spreads false information (e.g., misinformation or disinformation), rumors, fake news, or hoaxes [12]. Fake news is usually observed as more novel than real news; in addition, it flows on social/mass media noticeably faster, farther, and more broadly than real news [13]. Fake news has been commonly used to manipulate and propagate false information by appealing to users’ ideological perspectives, emotions, and desires to spread their views to other people [14]. Thus, the dissemination of fake news via social/mass media may have an effect on people’s social behavior. Social behavior changes can affect people’s well-being and resource distribution, resulting in changes in community resilience. However, prior studies have rarely assessed community resilience via social media and have rarely investigated the correlation between various types of news and tweets from the community resilience’s point of view.

### Research Goal, Contributions, and Questions

In this work, we aim to quantify community resilience (CR) in terms of community wellbeing (CW) and resource distribution (RD). These two factors are quantified by natural language processing (NLP) tools on news articles that include real, mixed (i.e., half fake and half real), and fake news as well as tweets including real and fake tweets. We also examine the correlation between the measured CR from news articles and the actual state of CR captured from tweets on Twitter.

In Fig. 1, we illustrate our proposed framework for measuring community resilience of various types of news/tweets using machine learning, natural language processing, and dimension reduction techniques.

**Fig 1.**
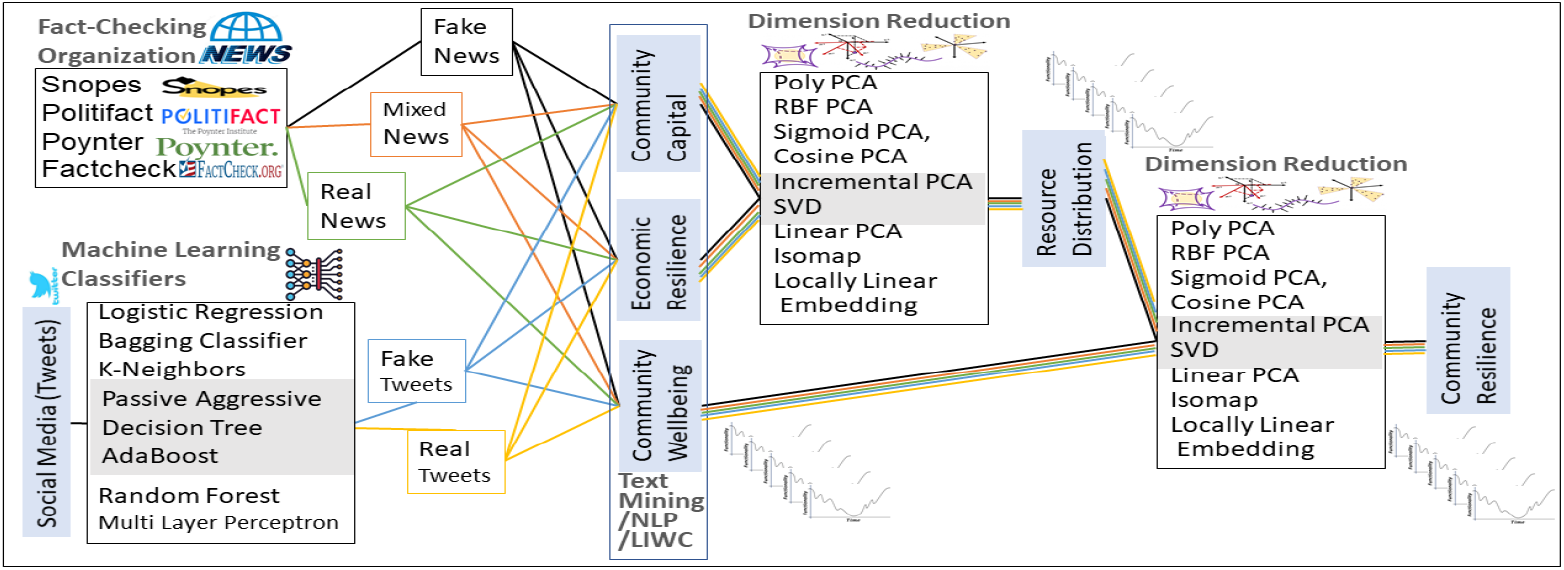
The proposed framework for assessing community resilience of various types of news/tweets via machine learning, natural language processing, and dimension reduction techniques.

The **key contributions** of this work are as follows:

1. We develop novel community resilience metrics inspired by the system resilience metric in the cybersecurity domain [15]. We define community resilience in terms of a community’s absorption (or fault tolerance) to, adaptability to, and recoverability from attacks or failures (e.g., disasters). Specifically, we measure community resilience based on two attributes, namely, community wellbeing and resource distribution. We measure community wellbeing based on mental and physical wellbeing. We estimate resource distribution based on economic resilience, and community capital. To the best of our knowledge, measuring CR based on social media information has been rarely studied.

2. This work is the first to use news articles and Twitter to assess community resilience during the COVID-19. We use fact-checking to collect 4,952 full-text news articles and categorize them as real, mixed, or fake news. In addition, we retrieve tweets from 42,877,312 tweets IDs from Jan. 2020 to Jun. 2021. We use the top three machine learning (ML) algorithms, i.e., Passive-Aggressive Classifier, Decision Tree Classifier, and AdaBoost Classifier, to identify if a tweet is real or fake.

3. To boost the sociological significance of this work, we use dimension reduction techniques, including linear transformations, nonlinear transformations, and manifold learning to integrate various dimensions of community resilience. We will show that while the incremental principal component analysis (PCA) keeps temporal dependency information, it has a greater level of variance information ratio.

4. We analyze the correlation between measurements of CR attributes by each type of news (i.e., real, mixed, or fake) and tweets (i.e., real or fake). From this analysis, fake news is shown to influence people’s behaviors towards undesirable states, undermining CR in reality. Moreover, the CR measured based on real or mixed news articles can reflect actual states of the CR measured from tweets.

5. We conduct a resilience analysis of various types of news (i.e., real, mixed, or fake) and tweets (i.e., real and fake) via an output-oriented analysis to show the values of each CR attribute over time, as well as a capacity-based analysis to demonstrate the time-averaged CR measurements. We also conduct statistical analyses to examine the correlation of CR attributes measured from news and tweets.

Our study will answer the following **research questions**:

1. *What are the main trends observed in community resilience and its key attributes, i*.*e*., *community wellbeing and resource distribution?*
2. *What are the key differences and correlations between the community resilience measured on various types of news and tweets?*
3. *What are the levels of the community resilience metrics, e*.*g*., *absorption and recovery during COVID-19 on various types of news and tweets?*

### Research Assumptions and Limitations

We conduct our study by assuming the following intuitions. First, real tweets/news can represent community resilience better than mixed/fake tweets/news. Second, knowing a current situation with accurate information can lead people to make more rational decisions to handle a faced disaster, which is COVID-19 in this work. Although the scope of this work is limited to measuring and analyzing community resilience using tweets and news, further investigation to prove the above as the hypothesis will be conducted in our future work. As no research work cannot be faultless, our work also has a number of limitations:

1. While we gather all real and fake news propagated by the media, we only use Twitter to investigate the population’s behavior. To analyze population behavior, surveys can provide high-quality data, albeit at a cost. While a national survey is beneficial, it is an expensive and time-consuming endeavor. Due to the fact that we wish to track multiple metrics of community resilience over an extended period of time, the availability of datasets is critical. Note that there is a trade-off between the quality, sample size, period, availability, and cost of datasets. Further research can compare the correlations between fake/real news and surveys.
2. We use anxiety, anger, and sadness to determine the level of community wellbeing. Additional wellbeing metrics can be added. This necessitates the development of new techniques for assessing other possible community well-being indicators.
3. While Twitter may not be representative of the US population, it can provide insight into how people live their lives. Nonetheless, considering additional social media platforms may be beneficial for future research.

### Related Work

*Community resilience* (CR) refers to the ability of a social system to absorb the impact of the stress and cope with threats and adapt to post-event situations by reorganizing, changing, or learning to cope with the threat from the disasters [16, 17]. This definition is well aligned with the general concept of system resilience in terms of its fault tolerance (i.e., functioning under threats or errors), adaptability (i.e., adapting to disruptions), and recoverability (i.e., recovering quickly from the disrupted situations) [15]. Community resilience has been measured based on various types of metrics [18, 19, 20]. CR can be defined differently depending on different disasters faced in the past [21]. However, it has been commonly considered with a measure of resilience whether a society functions in terms of social, economic, institutional, infrastructure, community capital, and ecological aspects [22, 23].

Work [24] proposed the *wellbeing theory* discussing a measure of community wellbeing in terms of positive emotions, engagement, relationships, meaning, and accomplishment. [1] discussed ‘health’ in terms of behavioral, physical, social, and environmental wellbeing. Higher psychological wellbeing can introduce higher sustainability, equality, resilience, and inclusion [1, 24]. The key factors impacting people’s resilience to disasters were also studied, such as family distress, available support systems, disruption of school/job programs, or loss of loved ones/property [25].

The distribution state of physical and social resources is another indicator of community resilience. Physical resources consist of critical infrastructures, electricity, water, food, medicine, emergency services capacity, transit capacity, grocery, pharmacy, or workplaces. Social resources include community capital and institutional resources [26], which allow people to interact with other people for their social activities. During the COVID-19, we observed aggressive panic buying behaviors of food, toilet papers, and sanitary products across countries or regions such as Singapore [27], Hong Kong [28], and Chinese mainland [29]. This is known to reduce community resilience due to a lack of balanced resource distribution.

Social media activities influence community resilience [30] in terms of social wellbeing and community capital. Official and informal sources use social media to spread information to handle a disaster for public safety, such as social distance, sanitation, food or transportation availability, or business hours. In addition, social media provide good networking tools to engage people with a community or government guidance [31]. However, false information has often been propagated through social media, such as fake news or rumors, which can easily amplify fear, anxiety [27, 32], outright racism, disgust, and mistrust [28]. These unnecessary misperceptions have been the key to triggering irrational, undesirable responses to disasters. In the literature, people’s responses and behaviors to the COVID-19 have been measured by analyzing social media information. The examples include emotions and psychological states extracted from the datasets of Weibo users using the linguistic inquiry, word count (LIWC) framework [33, 34], risk perception, negative emotions (e.g., sadness, anger, anxiety), and behavioral responses (e.g., panic buying) to COVID-19 from the dataset of Sina Weibo, Baidu search engine, and Ali e-commerce marketplace using LIWC [35]. Aggressive panic buying behaviors were more prominently observed when more misinformation or rumors on the COVID-19 were disseminated [35]. Emotions (e.g., surprise, disgust, fear, anger, sadness, anticipation, joy, and trust) in replies were also captured from real and false tweets using the National Research Council Canada (NRC) [36] and LIWC [13]. Mingxuan *et al*. [37] measured people’s mental health based on emotions extracted from social media data, which was analyzed using machine learning (ML) or NLP techniques [38, 39].

However, to our knowledge, no prior work has estimated community resilience based on community wellbeing and resource distribution using both social media news articles (i.e., real, mixed, and fake) and tweets (i.e., real and fake) to compare their measurements and investigate their correlations.

## Measurement of Community Resilience Using Social Media Information

In this section, we discuss how community resilience is measured using social media information, including both news articles and tweets.

### Community Resilience Metrics

We measure the community functionality in terms of community wellbeing and resource distribution. Fig. 2 represents the community functionality, *CF* (*t*), with time *t*. We define community resilience based on the concept of system resilience [15], consisting of absorption (i.e., fault tolerance), adaptability, and recoverability. We interpret the time until a community does not function as the time period for absorption, namely TFA (i.e., time from *t*_0_ to *t*_1_). Absorption (ABS) refers to the community’s capacity to absorb the shock and adverse effects caused by COVID-19. High TFA implies that the community tolerates hardships introduced by a disaster so that the community can still function by providing at least critical, minimum services, such as food, employment, schools, or health services. Note that a higher absorption is more desirable. Community Non-Functioning (CNF) is a term that refers to situations in which the community’s functionality falls below a critical threshold. We denote the deadlock functionality threshold by *b*. We call the time from *t*_1_ to *t*_3_ the *time under community non-functioning* (TNF). A shorter TNF is considered more desirable, representing fast failure and fast recovery. By following the conventional concept of system reliability, *the mean time to recovery* (MTTR), we defined the time to recovery (TTR) estimated from the time the community reaches a critical functionality point (*t*_1_) to the time it fully recovers from the disaster and reaches at the initial normal state (*t*_4_). Recovery (REF) refers to the community’s capacity to recover from COVID-19. The recoverability effectiveness (RE) refers to how much the community has recovered from the minimum functionality point, *t*_2_, to the current point at *t*_4_. Note that a higher level of recovery is more desirable. We consider the whole period from the outbreak of a disaster (e.g., COVID-19) to the time a community is fully recovered, *t*_4_, as the time period for adaptability (TA). Depending on how the community handles the disaster, TA may not face TNF but directly recover from a less functionality state to a full functionality state.

**Fig 2.**
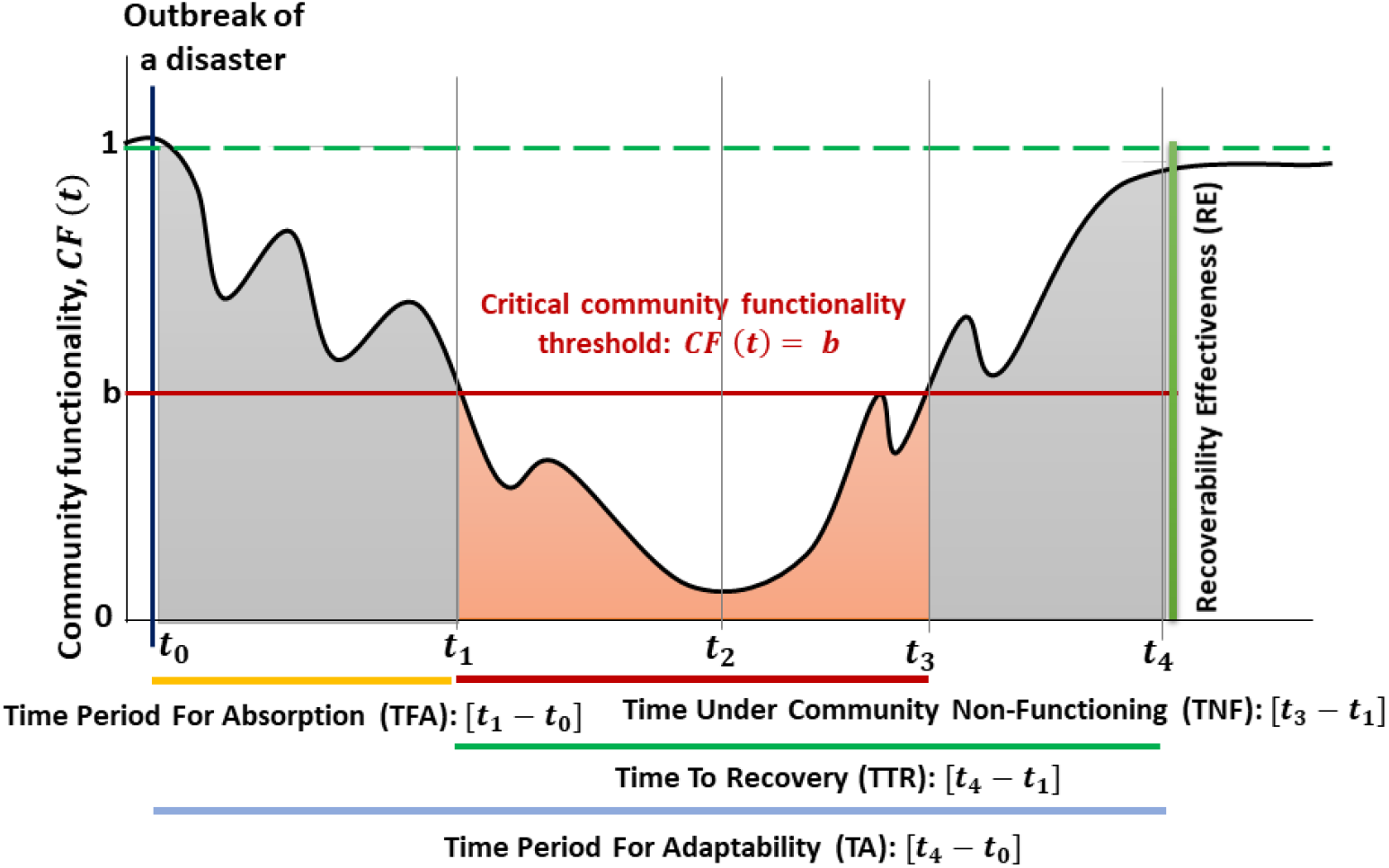
The evolution of community functionality (*CF* (*t*)) from the outbreak of a disaster (e.g., COVID-19) to the full recovery of a community.

Higher absorption, recovery, and adaptability are more desirable, which means the more area under the curve a community has, the more resilient it is.

We estimate *CF* (*t*) based on the levels of community wellbeing (*CW* (*t*)) and resource distribution (*RD*(*t*)) at time *t*. Here, *CR* is measured by:

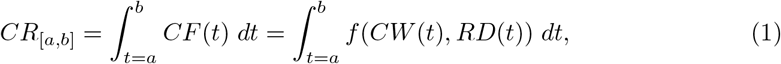

where [*a, b*] denotes the time period used to calculate *CR*. Note that *CW* and *RD* are treated equally in this work. For fair consideration of each component, we use a normalized value of *CW* and *RD* as a real number ranging in [0, 1] using *min-max scaling* [40]. Function *f* in its simple form can be the average of *CW* and *RD*. However, depending on the relative relevance of *CR* in a given domain, *CW* and *RD* can be weighted differently. In order to improve the operationality of this work, we will explore the appropriate *f* function. We will demonstrate that the incremental PCA function is the best *f* function.

To determine the average *CF* during the period of the COVID-19, we measure *ABS, CNF*, and *REF* as follows:

*CF* (*t*)

- *ABS* is the average *CF* during the time period for absorption, which is given by:

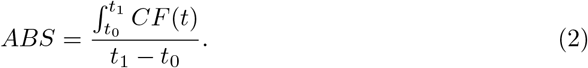
- *CNF* is the average *CF* over the time under the critical area of *CF*, which is measured by:

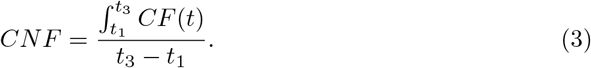 We assume that a community is entirely dysfunctional when its *CR* is below the threshold *b*.
- *REF* refers to the average *CF* during the period of recovery, which is obtained by:

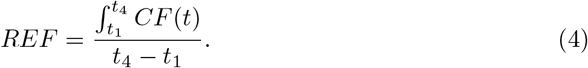

#### Integrating community resilience components

Since community resilience encompasses a variety of dimensions, the manner in which these characteristics are interwoven is critical. One strategy is to use a weighted average. To improve the operationalization and sociological significance of this work, we use dimension reduction techniques, including linear transformations, nonlinear transformations, and manifold learning to combine two main dimensions into one dimension, i.e., resource distribution or community resilience. We use multiple dimension techniques to determine which one performs better. Thus, the polynomial (Poly) Kernel PCA, the Gaussian radial basis function (RBF) Kernel PCA, the sigmoid Kernel PCA, the cosine Kernel PCA, the incremental PCA, the linear PCA, the SVD, the isomap, and the Locally Linear Embedding are the methods used to calculate resource distribution and community resilience. The variance information ratio (derived using the eigenvalues’ values), the reconstruction error, and the time-related correlations (time corr) are shown in Table 1. It is preferable to have a higher level of variance information ratio and a lower level of reconstruction error. While we are concerned with minimizing error, we also want to retain time-series information. In other words, this type of data is intrinsically associated with temporal dependency. As a result, we can determine the time-related correlation, or the correlation between the integrated result and each of its components. To be more precise, we calculate the correlation between resource distribution and each of community capital and economic resilience. Plus, we calculate the correlation between community resilience and each of community wellbeing and resource distribution. To maintain the temporal dependency information, at least one correlation should be positive. If two dimensions are raised, the integrated results should also increase. In Table 1, two techniques stand out, namely incremental PCA and SVD. Furthermore, based on the results of other techniques, there are scenarios in which there are two negative correlations. Because incremental PCA has a greater level of variance information ratio, we select it as the ultimate dimension reduction strategy.

**Table 1.**
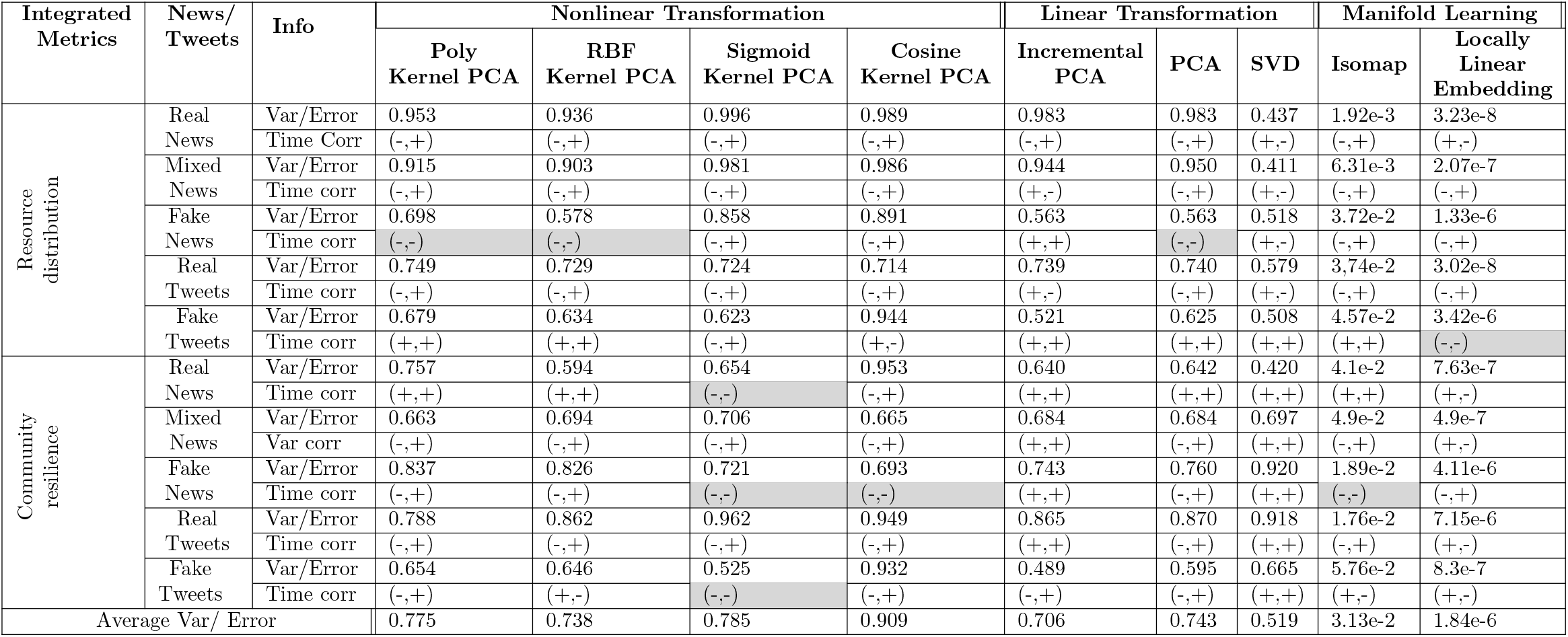
The variance information ratio (Var), the reconstruction error (error), and the time-related correlations (Time corr) of Resource distribution and community resilience by using the polynomial (Poly) Kernel PCA, the Gaussian radial basis function (RBF) Kernel PCA, the sigmoid Kernel PCA, the cosine Kernel PCA, the incremental PCA, the linear PCA, the SVD, the isomap, and the Locally Linear Embedding.

Now we describe how to estimate CW and RD as below.

#### Measuring Community Wellbeing

A lack of community wellbeing (CW) under disasters can either increase people’s vulnerability to early deaths or injuries, or trigger irrational behavior, such as panic buying [41]. Wellbeing is measured by the extent of people’s moods, such as anxiety, depression, and anger, which have long been recognized as typical symptoms of wellbeing illness [42, 43, 44]. Therefore, we obtain the extent of community wellbeing from the features of *anxiety, sadness*, and *anger*, extracted from linguistic inquiry and word count (LIWC) categories.

#### Measuring Resource Distribution

Resource distribution (RD) also measures part of CR [3, 4, 26] where the high functioning in RD refers to the high ability that a community can provide services to its inhabitants related to economic, infrastructure, institutional, and community capital resources. We assume that sufficient and well-distributed resources can contribute to the community that can better resist, recover, and/or overcome a disaster. We measure RD in terms of how well each service is provided. RD is measured by:

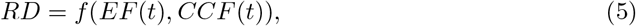

where *EF* (*t*) and *CCF* (*t*) refer to the level of states related to economic, and community capital functioning, respectively, with an equal weight considered. Again, depending on the domain requirement, its weight can be differently considered. As discussed before, function *f* can be as simple as the average of *EF* (*t*), and *CCF* (*t*). However, in order to improve the operationalization of this work, We will demonstrate that the incremental PCA function is the best *f* function.

Each component of RD, including *EF* (*t*), and *CCF* (*t*), is measured by LIWC categories as follows:

- *Economic functionality* (EF) is the economic capacity of a given community before and after a disaster. The examples include housing capital, employment, income, signal sector employment dependence, or business sizes. Economic functioning is captured by extracting the amount of words related to money or work, such as the increased use of work-related (e.g., ‘job,’ ‘majors,’ ‘xerox’), money-related (e.g., ‘Audit,’ ‘cash,’ or ‘owe’) terms in the LIWC categories.
- *Community capital* indicates a community’s ability to provide social activity services to its inhabitants and build trust among them. We assess community capital in terms of the language patterns representing community cooperation using the LIWC categories as follows:
  – *Communication Efficiency*: The increased use of complex words and words with more than six letters has been identified as being inefficient for communication and cooperation [45]. To measure this, we calculate the opposite degree of ‘Words*>* 6 letters.’
  – *Group-Oriented Communications*: The frequent use of first-person pronouns, such as ‘we,’ ‘us,’ ‘our,’ indicates group interaction [46]. In psychological linguistics, it is known that assent-related languages (e.g., ‘agree, ‘OK,’ ‘yes’) point to group consensus and cooperation [47]. Hence, we measure the frequency of words using the ‘first-person plural’ pronounces and ‘assent’ in the LIWC categories.
  – *Social Process-Related Communications*: We measure increased social engagement and cooperation [48, 49] based on the frequency of social process languages obtained by ‘friend’ and ‘family.’

The presence of more words within a category indicates a higher value. For fair comparison, we normalize the value of each attribute in CR by dividing the accumulated degree by the number of words, representing the extent of each attribute ranging in [0, 1] as a real number. Note that we can define community wellbeing, community capital, economic resilience, resource distribution, and community resilience in terms of absorption, adaptability, and recoverability components.

### Procedures of Measuring CR via Social Media Information

#### Collecting News Using Web-Scraping

We describe the process of finalizing information associated with news in Fig. 3. The information includes the text of news articles, issues, subjects, misconceptions, and the title of news articles for all the articles published over time. We use a two-stage web-scraping method to collect these contents. The web crawling process begins with the Google Chrome Extension ‘Web Scraper – Free Web-Scraping’ [50]. This tool allows interaction with the website from which we scrape data to identify the HTML tags required to extract data from fact-checking websites. We can export the results as a CSV file containing external links to the original articles. Then, we use the Python library Beautiful Soup [51] to analyze external links and scrap the original articles and additional tags that were difficult to web-scrape with the first tool. Additionally, we extract the quotation’s text from news scraped from fact-checking organizations. Then, we compare the *cosine similarity* [52] of this quoted text to the news obtained via external links to choose the most appropriate news text automatically and double-check them manually. Note that we filter the so-called ‘most appropriate news’ by capturing the original news text. The original news text is filtered out by excluding text quoted from other sources. We leverage the automatic web-scraping techniques to capture only the original news text solely written by the author of the given news article.

**Fig 3.**
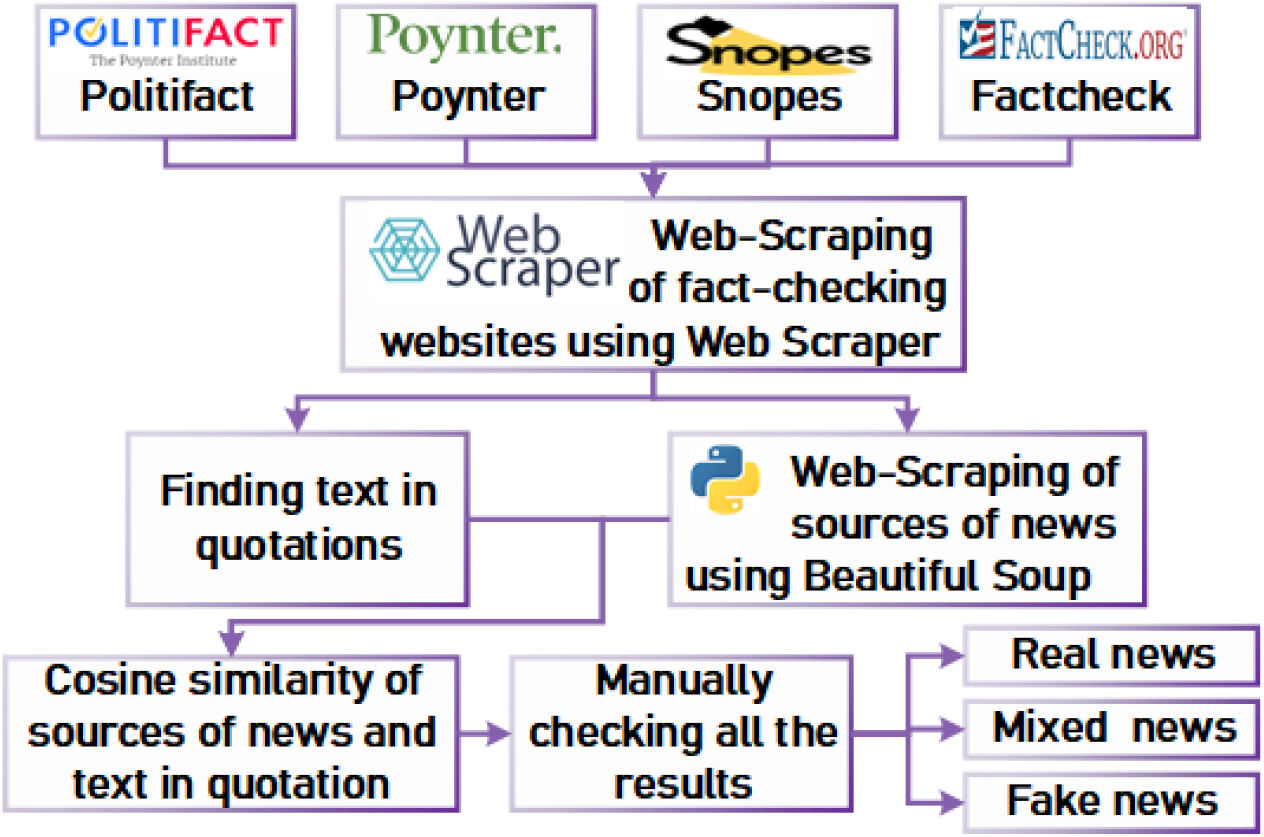
Collecting news based on web-scraping and manual cleaning.

#### Classification of News Articles

We extract 4,952 real, mixed, and fake news articles talking about COVID-19 based on the results of four fact-checking organizations, including Snopes [53], Politifact [54], Poynter [55], and Factcheck [56]. We gather 2413, 927, 1308, and 304 news articles talking about COVID-19 for Jan. 2020 - Jun. 2021 from these four organizations, respectively. It is not uncommon for fake news to be examined by several facts checking organizations. According to our datasets, no disagreement is found between these fact-checking outcomes across organizations. The categories of Snopes of interest include true, mostly true, mixture, mostly false, false news. Similarly, Politifact uses tag news with true, mostly true, half true, mostly false, false, and pants on fire news. We categorize news articles into real, mixed, or fake, as described in Table 2. Using these classifications, we collect all news articles from the archived news regarding COVID-19 from these organizations for Jan. 2020 - Jun. 2021.

**Table 2.** News types based on the classifications of three fact-checking organizations.

| Type of news | Snopes | Politifact | Poynter | Factcheck |
| --- | --- | --- | --- | --- |
| Real | true, mostly true | true, mostly true | - | - |
| Fake | mostly false, false | mostly false, false, pants on fire | fake | fake |
| Mixed | mixture | half true | - | - |
| Number | 2413 | 927 | 1308 | 304 |

#### Processing of News Articles for Analysis

We extract 3,437 news articles tagged with COVID-19 and coronavirus. After processing the initial cleaning, such as checking news with a correct tag, we come up with 3,235 news, consisting of 360 real news, 207 mixed news, and 2,668 fake news. After eliminating repetitive or irrelevant news, we select 207 news at random out of each pool of different types of news for fair consideration. Table 3 provides the distribution of published news and tweets considered across months. As in Table 3, we observe a significant amount of news articles published in Mar./Apr. 2020 and prominently there is a higher amount of fake news and tweets compared to those of real counterparts.

**Table 3.** The numbers of various types of news and tweets per month considered in this study.

| Year |  | 2020 |  |  |  |  |  |  |  |  |  |  |  | 2021 |  |  |  |  |  | All |
| --- | --- | --- | --- | --- | --- | --- | --- | --- | --- | --- | --- | --- | --- | --- | --- | --- | --- | --- | --- | --- |
| Month |  | Jan. | Feb. | Mar. | Apr. | May | Jun. | Jul. | Aug. | Sep. | Oct. | Nov. | Dec. | Jan. | Feb. | Mar. | Apr. | May | Jun. | All |
| News | Real | 0 | 3 | 35 | 47 | 27 | 18 | 28 | 16 | 12 | 15 | 8 | 32 | 27 | 13 | 23 | 25 | 21 | 10 | 360 |
|  | Mixed | 0 | 2 | 29 | 35 | 24 | 6 | 10 | 5 | 7 | 8 | 13 | 11 | 16 | 13 | 10 | 7 | 9 | 2 | 207 |
|  | Fake | 49 | 120 | 485 | 468 | 267 | 108 | 131 | 87 | 77 | 116 | 60 | 122 | 61 | 86 | 129 | 115 | 126 | 61 | 2668 |
|  | All | 49 | 125 | 549 | 550 | 318 | 132 | 169 | 108 | 96 | 139 | 81 | 165 | 104 | 112 | 162 | 147 | 156 | 73 | 3235 |
| tweets | Real | 124 | 81 | 316 | 208 | 138 | 91 | 46 | 40 | 29 | 35 | 20 | 196 | 114 | 88 | 87 | 93 | 83 | 76 | 1865 |
|  | Fake | 1993 | 1378 | 8463 | 5644 | 3391 | 1989 | 1364 | 904 | 720 | 894 | 559 | 3304 | 2716 | 2018 | 1762 | 1852 | 1854 | 1595 | 42400 |
|  | All | 2117 | 1459 | 8779 | 5852 | 3529 | 2080 | 1410 | 944 | 749 | 929 | 579 | 3500 | 2830 | 2106 | 1849 | 1945 | 1937 | 1671 | 44265 |

The news sources are mainly newspaper interviews, TV interviews, viral images, Journals, Press releases, digital ads, campaign ads, meeting in white houses, Story, TV segments, social media, or press conferences. The news is in the format of photos, infographics, videos, text, or interviews. As photos, infographics, videos, or interviews are not in the format of text, there is a challenge to analyze them. The fact-checking organizations put text and explanations related to each of them. Hence, we use the text generated by the fact-checking organizations to analyze them. We also use the converted format of the photo, infographic, or video for our analysis. We use the release date of the news to determine when a news article is published. The fact-checking organizations (i.e., Snopes, Politifact, Poynter, and Factcheck) categorize news into various classes based on Table 2.

#### Collecting COVID-19-Related Tweets

Twitter, one of the most famous platforms, has above 313 million active users who generate 500 million tweets per day [57, 58]. Hence, we investigated 42,877,312 tweet IDs for Jan. 2020 – Jun. 2021. Note that we limited tweets to the US and we ended up with 44,265 tweets. Furthermore, we ordered these tweets chronologically, as in Table 3, showing a significant amount of tweets generated during Mar./Apr. 2020.

#### Classifying All Tweets as Real or Fake Based on Three machine learning (ML) Classifiers

We first classify tweets as real or fake. We first train eight existing ML classifiers on the datasets described in [59], which contain 23,481 fake tweets and 21,417 real news articles. We then select the top three ML classifiers, i.e., Passive-Aggressive, Decision Tree, and AdaBoost based on their prediction performance, as shown in Table 4. Finally, we predict the truthfulness of each tweet using these three ML algorithms and determine the final prediction for each tweet based on the majority rule of the three ML classifiers (i.e., at least two ML classifiers should give the same prediction result).

**Table 4.** Prediction Performance of Various Machine Learning Classifiers.

| ML Classifier | Accuracy | Precision | Recall | F-score |
| --- | --- | --- | --- | --- |
| Passive Aggressive | 0.995 | 0.995 | 0.995 | 0.995 |
| Logistic regression | 0.984 | 0.984 | 0.984 | 0.984 |
| Bagging Classifier | 0.618 | 0.779 | 0.598 | 0.532 |
| K-Neighbors | 0.671 | 0.782 | 0.655 | 0.622 |
| Decision Tree | 0.994 | 0.994 | 0.994 | 0.994 |
| Random Forest | 0.519 | 0.623 | 0.5 | 0.346 |
| AdaBoost | 0.995 | 0.995 | 0.995 | 0.995 |
| Multi Layer Perceptron | 0.966 | 0.967 | 0.966 | 0.966 |

#### Identifying Physical-Psycho-Social States and Behavioral Patterns using LIWC

We use the LIWC as our text-mining tool for the analyses of COVID-19 related news and tweets because it contains a wealth of physical-psychosocial characteristics and behavioral patterns. Prior to analyzing them with the LIWC, all tweets are sorted by month and cleaned using various NLP tools (i.e., nltk, string, stopwords, RegexpTokenizer, and regexp) for each type of news (i.e., real, mixed, or fake) and tweet (i.e., real or fake). We begin text cleaning by removing HTML, punctuation, stop words, and stammering words. Following that, we extract all LIWC features relevant to CR assessment.

## Experimental Results & Analysis

### News Analyses

Fig. 4 illustrates the word cloud associated with real, mixed, fake, and all news. Fig. 5 plots the positive and negative sentiments associated with various types of news over time. Table 5 shows the frequency of various topics under different types of news. Politics is the most popular subject. Medical and health, entertainment, and business are also popular topics affecting community resilience. In May and Sep. 2020, real news has the least positive and negative sentiment. In Sep. 2020 and Mar. 2020, mixed news has the least positive and negative content. In Jan. 2021 and Jun. 2020, fake news has the least positive and negative sentiment. In Sep. and Mar. 2020, all news is at its least positive and least negative, respectively. The subject of each news item is determined by fact-checking organizations, such as Snopes and Politifact.

**Table 5.** Frequency of Different Types of News Collected Under Various Topics for Jan. 2020 – Jun. 2021.

| Source | Subject (an amount of news) |
| --- | --- |
| Real news | politics (87), medical (29), fauxtography (17), entertainment (13), business (12), viral (5), phenomena (5), crime (5), history (5), health (5) |
| Mixed news | coronavirus (67), politics (48), health (32), facebook (29), public (19), medical (17), fact (16), checks (16), posts (10), budget (8) |
| Fake news | politics (97), medical (39), fauxtography (13), entertainment (9), junk (9), news (9), viral (7), phenomena (7), technology (6), business (5) |
| All news | politics (229), medical (84), coronavirus (67), health (38), fauxtography (31), facebook (29), entertainment (23), business (22), public (17), fact (16) |

**Fig 4.**
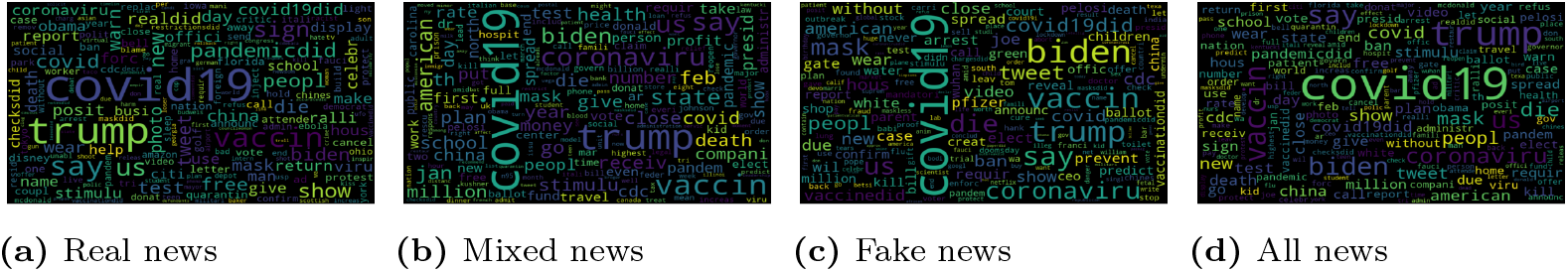
Word cloud for real, mixed, fake, and all news for Jan. 2020 – Jun. 2021.

**Fig 5.**
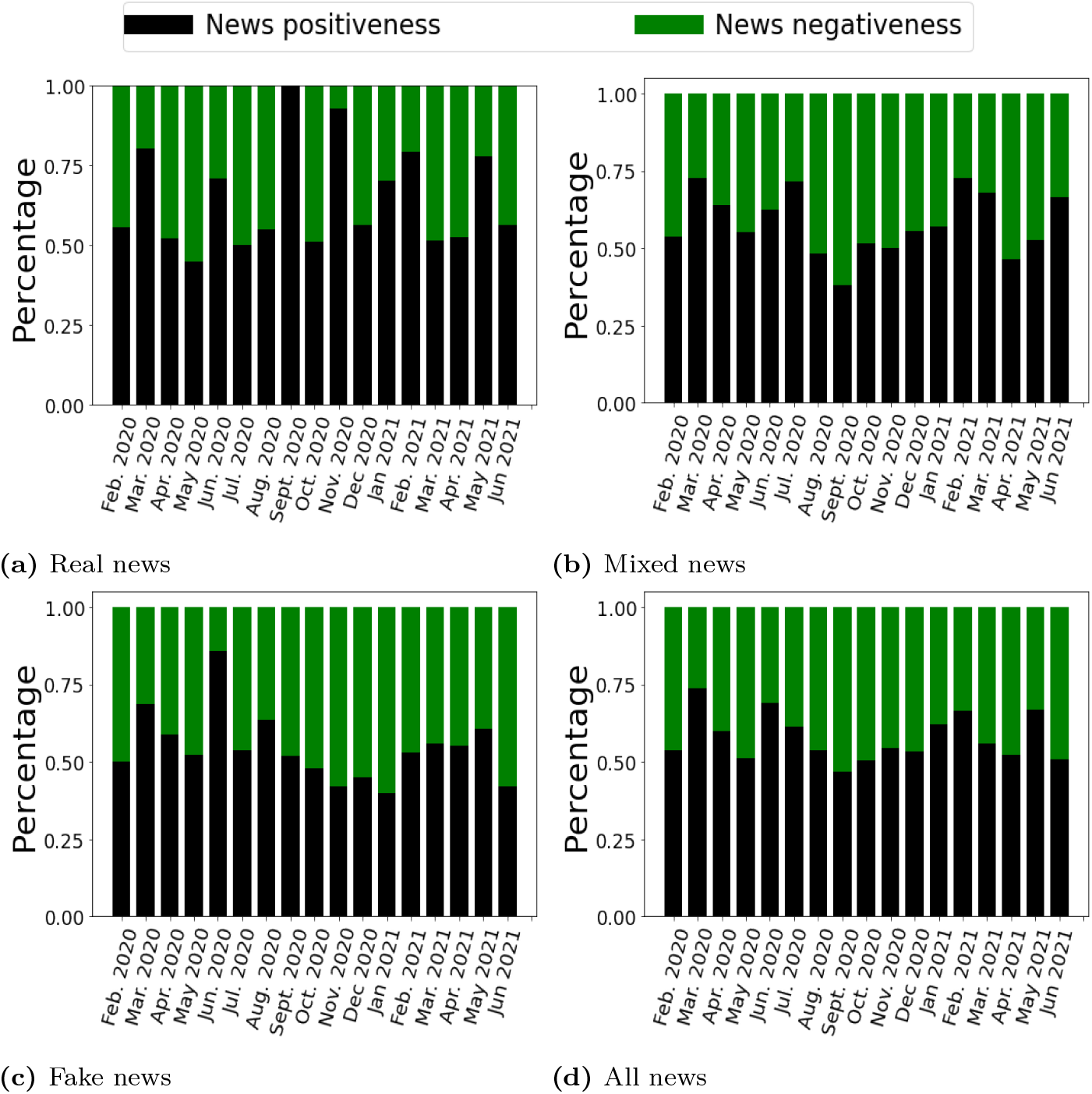
The positiveness and negativeness of news about the COVID-19 for Jan. 2020 – Jun. 2021.

### Community Wellbeing Assessment

The output-oriented analysis measurements provide accurate information about the trend and dynamic change of functionality in a given community [60]. From Feb. 2020 to Jun. 2021, Fig. 6 depicts the normalized degree of output-oriented community wellbeing (CW) as measured by real, mixed, and fake news as well as real and fake tweets. Fake news and fake tweets demonstrate similar CW patterns. The peak of CW in fake tweets and real/fake news occurs in Sep. 2020. On the other hand, the peaks of CW in real tweets and mixed news occur in Feb. 2020 and Jun. 2021, respectively. We also observe that CW reaches its lowest point by the end of 2020 under real tweets. This result aligns well with the trends reported by the US Census Bureau [61] that since the COVID-19 outbreak in Feb. 2020, people’s wellbing had deteriorated by the end of 2020.

**Fig 6.**
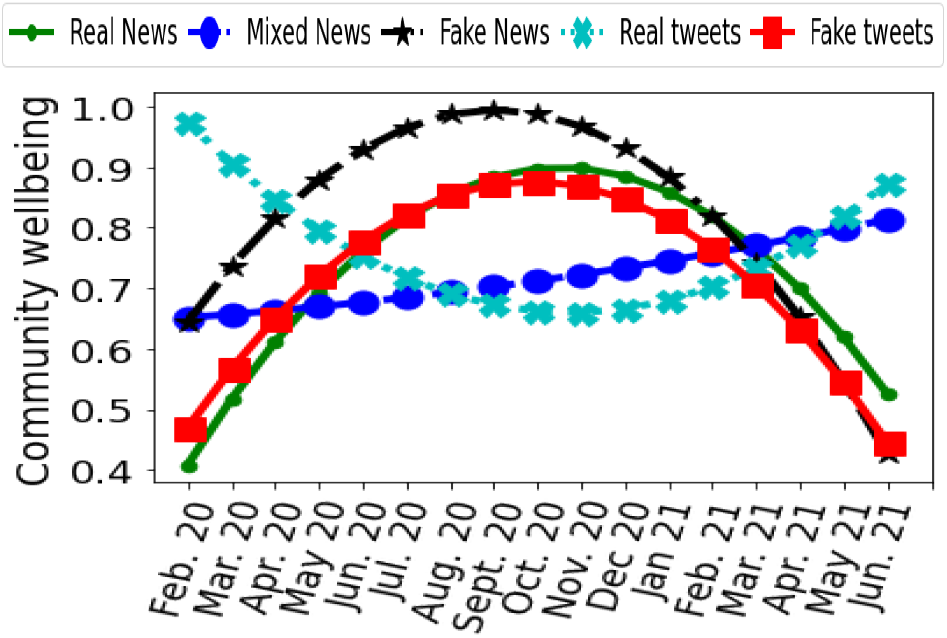
Community wellbeing measured by different types of news (i.e., real, mixed, and fake) and tweets (i.e., real and fake).

### Community Capital, Economic Resilience, and Resource Distribution Assessment

Fig. 7 illustrates the output-oriented degree of community capital, economic resilience, and resource distribution measured from the news (i.e., real, mixed, and fake) and tweets (i.e., real and fake) collected for Feb. 2020 – Jun. 2021. From this figure, we observe that real tweets and real news typically follow similar trends for Community capital, and economic resilience. Similarly, fake tweets and fake news also exhibit approximately similar trends.

**Fig 7.**
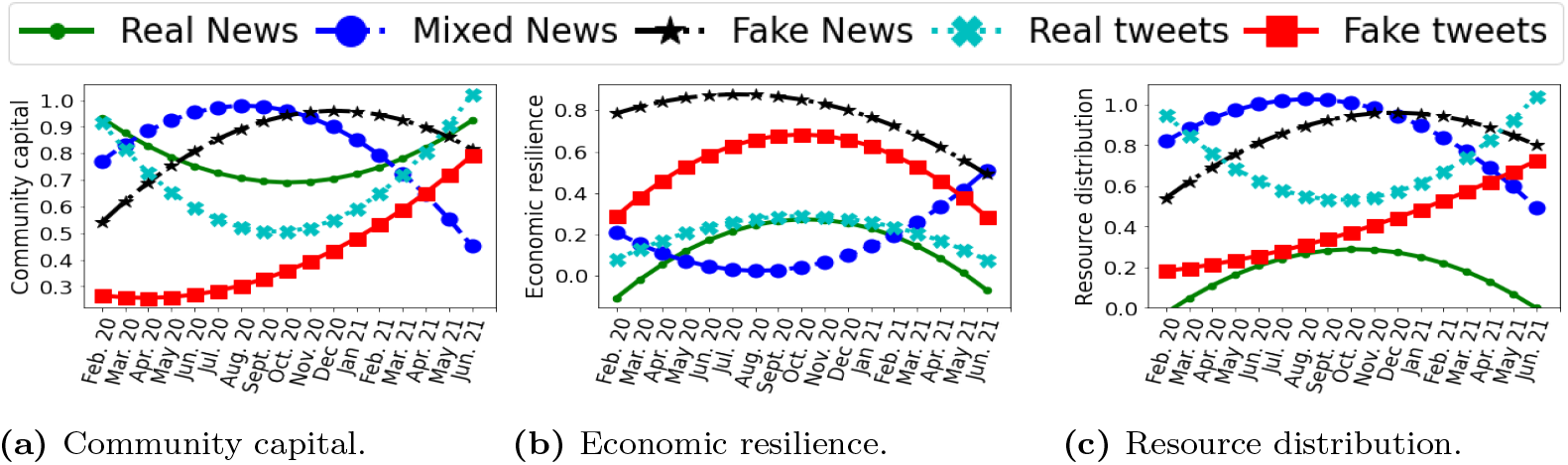
Community capital, economic resilience, and resource distribution measured based on different types of news and tweets for Feb. 2020–Jun. 2021.

For real news and tweets, economic functionalities are at their peak in Sep. 2020, while community capital is at the lowest level. Community capital shows its trend in the opposite direction of economic functionality for real/mixed news and real/fake tweets. This is because when a community is threatened due to the impact introduced by a disaster, people are more likely to cooperate for survival.

The incremental PCA method calculates the resource distribution based on community capital and economic resilience. The findings indicate that the trends in mixed/fake news and real/fake tweets are comparable to those in community capital. On the other hand, the trend in real news about resource distribution tracks the economic functionality trend. Fake tweets and fake news both exhibit the same pattern in terms of resource distribution. Simultaneously, real and mixed news follow similar trends.

### Community Resilience Assessment

#### Output-Oriented Resilience Assessment

We first measure CR over time (i.e., Feb. 2020 to Jun. 2021) for the output-oriented resilience assessment, as shown in Fig. 8. Although real news shows that community resilience begins to improve by the end of 2020, it also begins to deteriorate in 2021. In 2021, people’s wellbeing has been worsened. This is probably because people become tired of long-term restrictions in their daily lives, such as social distancing and online schooling/working, especially with the emergence of COVID-19 variants. These factors may drive people to become more pessimistic about the full recovery from the pandemic.

**Fig 8.**
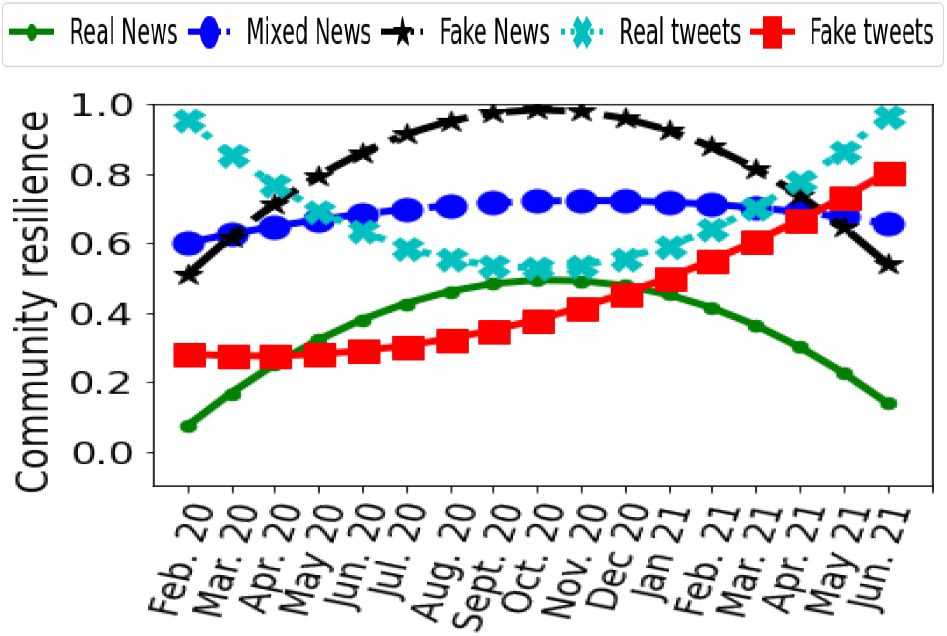
Output-oriented analysis of community resilience measured based on different types of news and tweets for Feb.2020–Jun. 2021.

Note that the incremental PCA method calculates the community resilience based on community wellbeing and resource distribution. The findings indicate that the trends in real/fake news and real/fake tweets are comparable to those in resource distribution. On the other hand, the trends in real, mixed, and fake news about resource distribution track the community wellbeing trends. Note that resource distribution and community wellbeing follow the same pattern for real and fake news.

#### Capacity-based Resilience Assessment

Capacity-based measurements are time-averaged community resilience (CR) measurements of a given community, indicating the degree of functionality of the community [60]. Fig. 9 illustrates the capacity-based values of all resilience-related metrics, including community wellbeing, community capital, economic resilience, resource distribution, and finally, community resilience, measured using real, mixed, and fake news as well as real and fake tweets.

**Fig 9.**
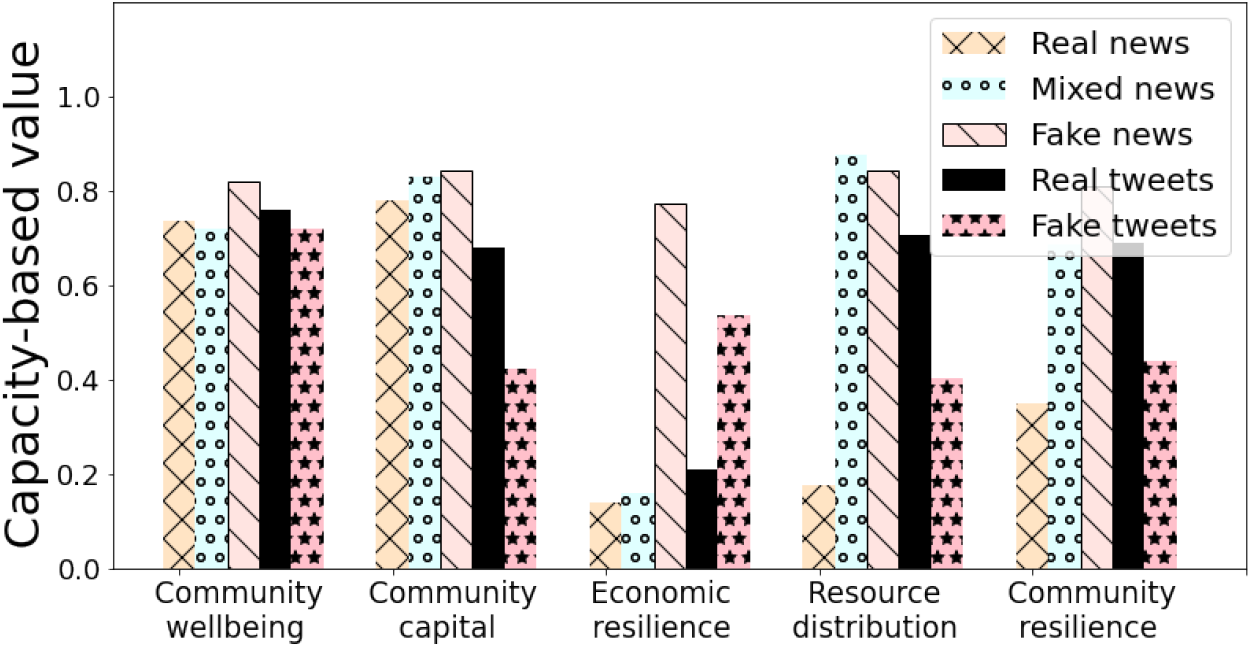
Capacity-based analysis of community wellbeing, community capital, economic resilience, resource distribution, and community resilience.

We observe from Fig. 9 that fake news is in a better state of community wellbeing (CW). In other words, released fake news implies that CW is adequate and likely underestimates the detrimental effect of the COVID-19. Additionally, people’s communication via fake tweets demonstrates a significant level of isolation, whereas real tweets show a higher level of community capital. Fig. 9 shows that while fake news presents a high degree of economic resilience, real news shows a low degree of economic resilience under the COVID-19. A possible reason is that fake news can trigger panic buying, thus eroding economic resilience. Similarly, fake news has a greater level of resource distribution than real news. Finally, fake news shows higher CR than real news. Fake news has the potential to mislead people into taking inappropriate actions in response to the COVID-19 by forming unrealistic optimism about the future. For instance, some fake news suggests that smoking, self-medicating with antibiotics, and wearing multiple surgical masks help combat COVID-19. This information is not only impractical, but also potentially jeopardizing community resilience.

### Absorption, Community Non-Functioning, and Recovery

Table 6 shows the measurement values of community functionality (CF) metrics, including Absorption (ABS), Community Non-Functioning (CNF), Recovery (REC), Time for Absorption (TFA), Time under Community Non-Functioning (TNF), and Time To Recovery (TTR) (see Fig. 2) for news and tweets, with the critical CF threshold, *b*, varying in the range of 0.2 to 0.5 in increment of 0.1.

**Table 6.** Absorption (ABS), Community Non-Functioning (CNF), Recovery (REC), Time for Absorption (TFA), Time under Community Non-Functioning (TNF), and Time To Recovery (TTR) for News and tweets with the Critical Community Functionality Threshold, *b*, varying over the range of 0.2-0.5.

| Adaptability Indexes | $b = 0.2$ | | | | | $b = 0.3$ | | | | | $b = 0.4$ | | | | | $b = 0.5$ | | | | |
| --- | --- | --- | --- | --- | --- | --- | --- | --- | --- | --- | --- | --- | --- | --- | --- | --- | --- | --- | --- | --- |
|  | News |  |  | tweets |  | News |  |  | tweets |  | News |  |  | tweets |  | News |  |  | tweets |  |
|  | Real | Mixed | Fake | Real | Fake | Real | Mixed | Fake | Real | Fake | Real | Mixed | Fake | Real | Fake | Real | Mixed | Fake | Real | Fake |
| **ABS** | 0 | 0.6 | 0.51 | 0.68 | 0.28 | 0 | 0.6 | 0.51 | 0.68 | 0 | 0 | 0.6 | 0.51 | 0.68 | 0 | 0 | 0.6 | 0.51 | 0.68 | 0 |
| **CNF** | 0.13 | 0 | 0 | 0 | 0 | 0.17 | 0 | 0 | 0 | 0.28 | 0.25 | 0 | 0 | 0 | 0.31 | 0.35 | 0 | 0 | 0 | 0.34 |
| **REC** | 0.35 | 0.69 | 0.81 | 0.68 | 0.46 | 0.35 | 0.69 | 0.81 | 0.68 | 0.44 | 0.35 | 0.69 | 0.81 | 0.68 | 0.44 | 0.35 | 0.69 | 0.81 | 0.68 | 0.44 |
| **TFA** | 0 | 1 | 1 | 9 | 3 | 0 | 1 | 1 | 9 | 0 | 0 | 1 | 1 | 9 | 0 | 0 | 1 | 1 | 9 | 0 |
| **TNF** | 3 | 0 | 0 | 0 | 0 | 5 | 0 | 0 | 0 | 5 | 9 | 0 | 0 | 0 | 9 | 17 | 0 | 0 | 0 | 12 |
| **TTR** | 17 | 17 | 17 | 9 | 15 | 17 | 17 | 17 | 9 | 17 | 17 | 17 | 17 | 9 | 17 | 17 | 17 | 17 | 9 | 17 |

Note that TFA, TNF, and TTR refer to the month-based average values.

Fake news induces a higher level of absorption for all critical CF threshold values than real news. Additionally, fake news typically exhibits the greatest degree of recovery. Fake news fosters distrust among the public, despite the fact that trust is a critical component of transparent risk communication, collaboration, and the cooperation of individuals to overcome catastrophic events. The negative outputs of fake news create problems not only in handling COVID-19 but also in recovering from it.

Real news induces a 17-month recovery for all critical CF threshold values, while the absorption level is 0-1 month. This means that CR steadily increased from Feb. 2020 to Jun. 2021. In other words, with real news, the community can recover very quickly following the initial degradation of functionality. Additionally, the number of months during which the community is non-functioning ranges from 0 to 17 months, depending on the critical threshold level. For example, TNF is equal to 17 months when *b* = 0.5 for real news, which means that the community functionality from the perspective of real news is less than 0.5 for all 17 months. Understandably, as the critical threshold level increases, the time duration associated with community dysfunction and recovery increases, while that associated with absorption decreases. On the other hand, mixed news has a higher level of absorption than fake news. Both fake news and mixed news show a higher level of absorption than that of real news. This implies that the level of community functionality is initially high and gradually declines, whereas real news demonstrates a rapid decline in community functionality at the start. Therefore, we can conclude that mixed/fake news tends to underestimate the negative impact of COVID-19 on the community. Real tweets, on the other hand, exhibit a high absorption level when *b* = 0.2 *−* 0.5, indicating that individuals believe the community is highly functional.

Tables 7-10 show the measurement values of community wellbeing, resource distribution, community capital, and economic functionality metrics, including Absorption (ABS), Community Non-Functioning (CNF), Recovery (REC), Time for Absorption (TFA), Time under Community Non-Functioning (TNF), and Time To Recovery (TTR), respectively. Based on the results:

**Table 7.** Absorption (ABS), Community Non-Functioning (CNF), Recovery (REC), Time for Absorption (TFA), Time under Community Non-Functioning (TNF), and Time To Recovery (TTR) for News and tweets with the Critical wellbeing Functionality Threshold, *b*, varying over the range of 0.2-0.5.

| Adaptability Indexes | $b = 0.2$ | | | | | $b = 0.3$ | | | | | $b = 0.4$ | | | | | $b = 0.5$ | | | | |
| --- | --- | --- | --- | --- | --- | --- | --- | --- | --- | --- | --- | --- | --- | --- | --- | --- | --- | --- | --- | --- |
|  | News |  |  | tweets |  | News |  |  | tweets |  | News |  |  | tweets |  | News |  |  | tweets |  |
|  | Real | Mixed | Fake | Real | Fake | Real | Mixed | Fake | Real | Fake | Real | Mixed | Fake | Real | Fake | Real | Mixed | Fake | Real | Fake |
| **ABS** | 0.41 | 0.65 | 0.82 | 0.77 | 0.72 | 0.41 | 0.65 | 0.82 | 0.77 | 0.72 | 0.41 | 0.65 | 0.82 | 0.77 | 0.72 | 0 | 0.65 | 0.84 | 0.77 | 0 |
| **CNF** | 0 | 0 | 0 | 0 | 0 | 0 | 0 | 0 | 0 | 0 | 0 | 0 | 0 | 0 | 0 | 0.41 | 0 | 0.43 | 0 | 0.46 |
| **REC** | 0.74 | 0.72 | 0.43 | 0.74 | 0.44 | 0.74 | 0.72 | 0.43 | 0.74 | 0.44 | 0.74 | 0.72 | 0.43 | 0.74 | 0.44 | 0.74 | 0.72 | 0.43 | 0.74 | 0.72 |
| **TFA** | 1 | 1 | 17 | 10 | 17 | 1 | 1 | 17 | 10 | 17 | 1 | 1 | 17 | 10 | 17 | 0 | 1 | 16 | 10 | 0 |
| **TNF** | 0 | 0 | 0 | 0 | 0 | 0 | 0 | 0 | 0 | 0 | 0 | 0 | 0 | 0 | 0 | 1 | 0 | 1 | 0 | 2 |
| **TTR** | 17 | 17 | 1 | 8 | 1 | 17 | 17 | 1 | 8 | 1 | 17 | 17 | 1 | 8 | 1 | 17 | 17 | 1 | 8 | 17 |

Note that TFA, TNF, and TTR refer to the month-based average values.

**Table 8.**
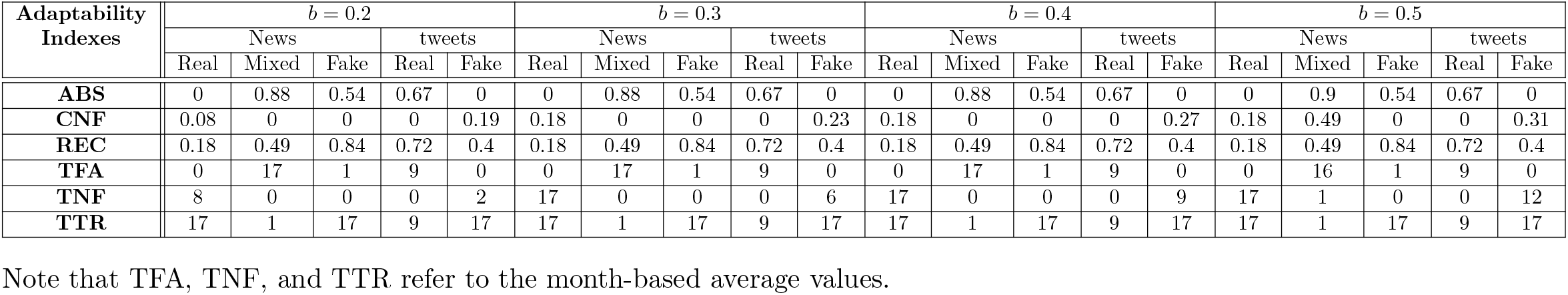
Absorption (ABS), Community Non-Functioning (CNF), Recovery (REC), Time for Absorption (TFA), Time under Community Non-Functioning (TNF), and Time To Recovery (TTR) for News and tweets with the Critical resource distribution Functionality Threshold, *b*, varying over the range of 0.2-0.5.

**Table 9.**
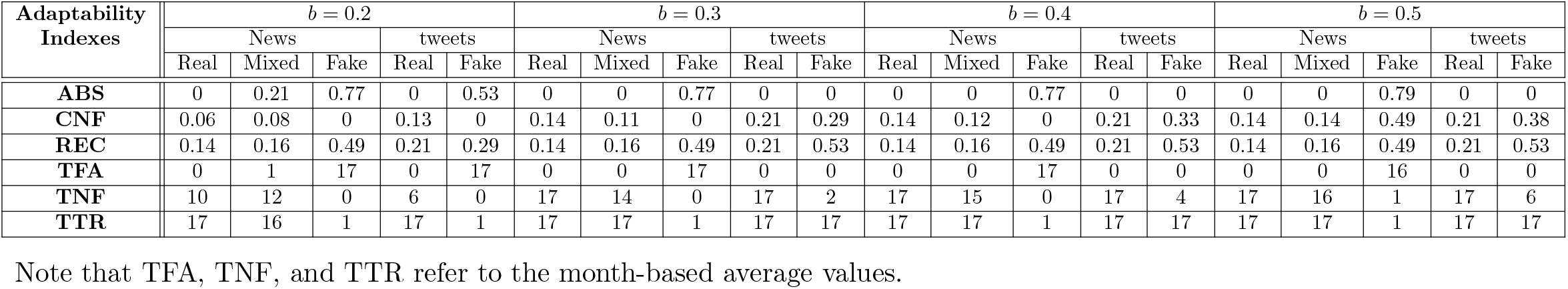
Absorption (ABS), Community Non-Functioning (CNF), Recovery (REC), Time for Absorption (TFA), Time under Community Non-Functioning (TNF), and Time To Recovery (TTR) for News and tweets with the Critical economic Functionality Threshold, *b*, varying over the range of 0.2-0.5.

| Adaptability Indexes | $b = 0.2$ | | | | | $b = 0.3$ | | | | | $b = 0.4$ | | | | | $b = 0.5$ | | | | |
| --- | --- | --- | --- | --- | --- | --- | --- | --- | --- | --- | --- | --- | --- | --- | --- | --- | --- | --- | --- | --- |
|  | News |  |  | tweets |  | News |  |  | tweets |  | News |  |  | tweets |  | News |  |  | tweets |  |
|  | Real | Mixed | Fake | Real | Fake | Real | Mixed | Fake | Real | Fake | Real | Mixed | Fake | Real | Fake | Real | Mixed | Fake | Real | Fake |
| **ABS** | 0 | 0.21 | 0.77 | 0 | 0.53 | 0 | 0 | 0.77 | 0 | 0 | 0 | 0 | 0.77 | 0 | 0 | 0 | 0 | 0.79 | 0 | 0 |
| **CNF** | 0.06 | 0.08 | 0 | 0.13 | 0 | 0.14 | 0.11 | 0 | 0.21 | 0.29 | 0.14 | 0.12 | 0 | 0.21 | 0.33 | 0.14 | 0.14 | 0.49 | 0.21 | 0.38 |
| **REC** | 0.14 | 0.16 | 0.49 | 0.21 | 0.29 | 0.14 | 0.16 | 0.49 | 0.21 | 0.53 | 0.14 | 0.16 | 0.49 | 0.21 | 0.53 | 0.14 | 0.16 | 0.49 | 0.21 | 0.53 |
| **TFA** | 0 | 1 | 17 | 0 | 17 | 0 | 0 | 17 | 0 | 0 | 0 | 0 | 17 | 0 | 0 | 0 | 0 | 16 | 0 | 0 |
| **TNF** | 10 | 12 | 0 | 6 | 0 | 17 | 14 | 0 | 17 | 2 | 17 | 15 | 0 | 17 | 4 | 17 | 16 | 1 | 17 | 6 |
| **TTR** | 17 | 16 | 1 | 17 | 1 | 17 | 17 | 1 | 17 | 17 | 17 | 17 | 1 | 17 | 17 | 17 | 17 | 1 | 17 | 17 |

Note that TFA, TNF, and TTR refer to the month-based average values.

**Table 10.**
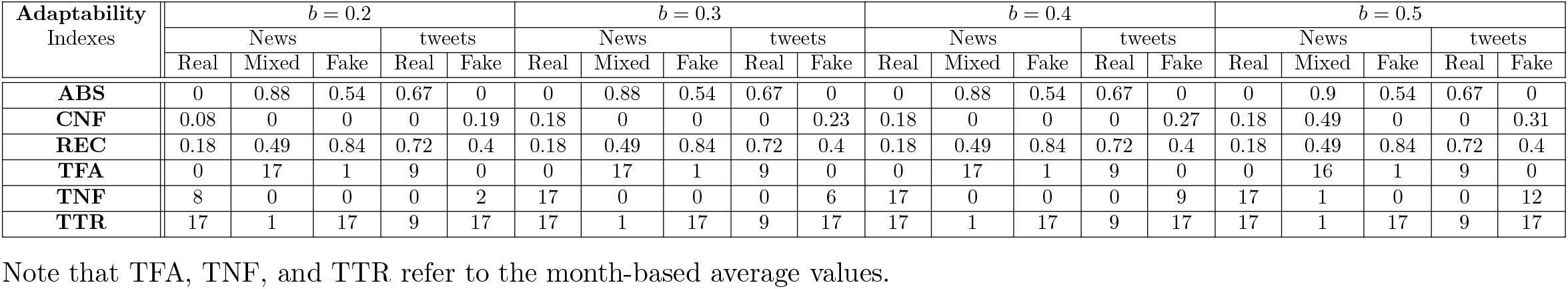
Absorption (ABS), Community Non-Functioning (CNF), Recovery (REC), Time for Absorption (TFA), Time under Community Non-Functioning (TNF), and Time To Recovery (TTR) for News and tweets with the Critical community capital Functionality Threshold, *b*, varying over the range of 0.2-0.5.

- Community wellbeing’s point of view: While fake news induces the highest level of absorption for all critical CF threshold values, real news typically exhibits the greatest degree of recovery.
- Resource distribution’s point of view: While fake news induces a higher level of absorption for all critical CF threshold values compared to real news, mixed news has the highest level of absorption. Additionally, fake news typically exhibits the greatest degree of recovery.
- Economic functionality’s point of view: Fake news induces the highest level of absorption and recovery for all critical CF threshold values,
- Community capital’s point of view: It is similar to Resource distribution.

### Statistical Analyses of News and Tweets

Table 11 shows the findings from our statistical analyses on the correlation between news and tweets. The statistical analyses include Pearson correlation (PC), Kendall tau correlation (KC), parametric statistical hypothesis tests (PT; Student’s t-test), and non-parametric statistical hypothesis tests (NT; Mann-Whitney U Test). The Pearson correlation (PC) and Kendall tau correlation coefficients demonstrate the linear and monotonic relationships between two variables, *x* and *y* [62]. We choose Pearson’s correlation coefficient to investigate if there is a linear statistical relationship or association between a resilience metric measured from real/mixed/fake news (*x*) vs. the same resilience metric measured from real/fake Tweets (*y*). The Pearson correlation coefficient assumes that both *x* and *y* are normally distributed. When this assumption does not hold, we rely on a non-parametric approach, such as Kendall tau correlation, which does not make any assumption about distribution. According to Table 11, fake tweets and news have a positive correlation for resilience-related features with a probability of 80 percent. Pearson and Kendall tau correlations (PC and KC) indicate that the correlations between fake news and real tweets are negative, with a probability of 80 percent. We also found that mixed news negatively correlates with real and fake tweets across all types of CR attributes with a probability of 95 percent. Parametric and non-parametric statistical hypothesis tests (PT and NT) demonstrate the distribution’s similarity across multiple scenarios. Fig. 10 illustrates the Quantile-Quantile (Q-Q)-plot for community resilience in relation to various news types (i.e., real, mixed, or fake) and tweet types (i.e., real or fake). We observe that fake tweets and real tweets exhibit similarity in their distributions with the probability of 60 percent. This similarity implies that both tweets can properly reflect the actual states of community resilience (CR) regardless of their truthfulness. Furthermore, analyzing social media information and predicting CR can provide a useful indicator to measure how our community is functioning against a disaster such as COVID-19.

**Table 11.**
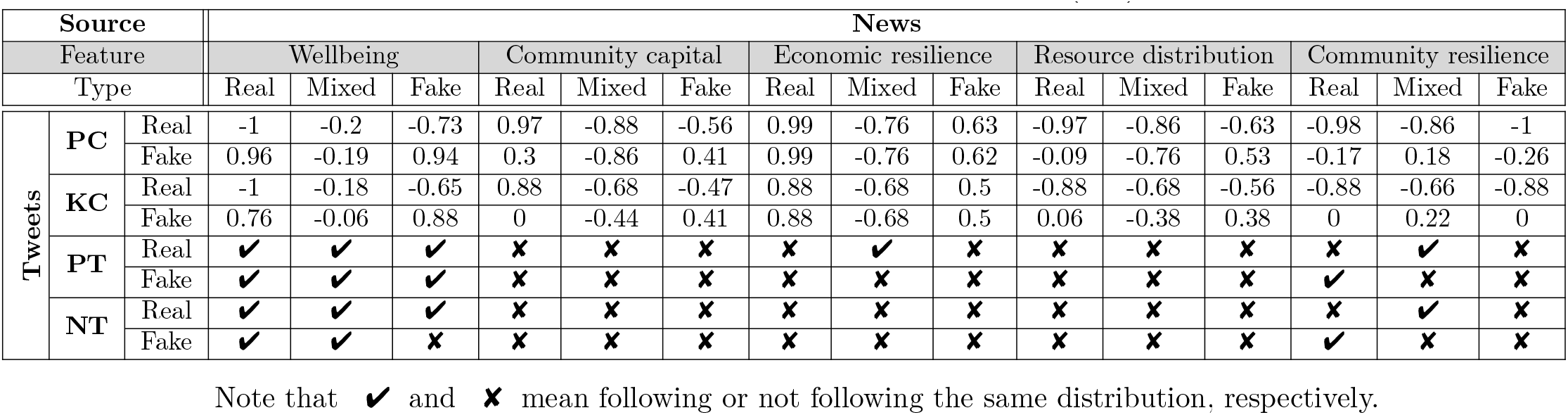
The Statistical Analysis of Various Functionalities for Three News Compared To Two Types of tweets: Pearson Correlation (PC), Kendall Tau Correlation (KC), Parametric Statistical Hypothesis Tests (PT), and Non-Parametric Statistical Hypothesis Tests (NT)

**Fig 10.**
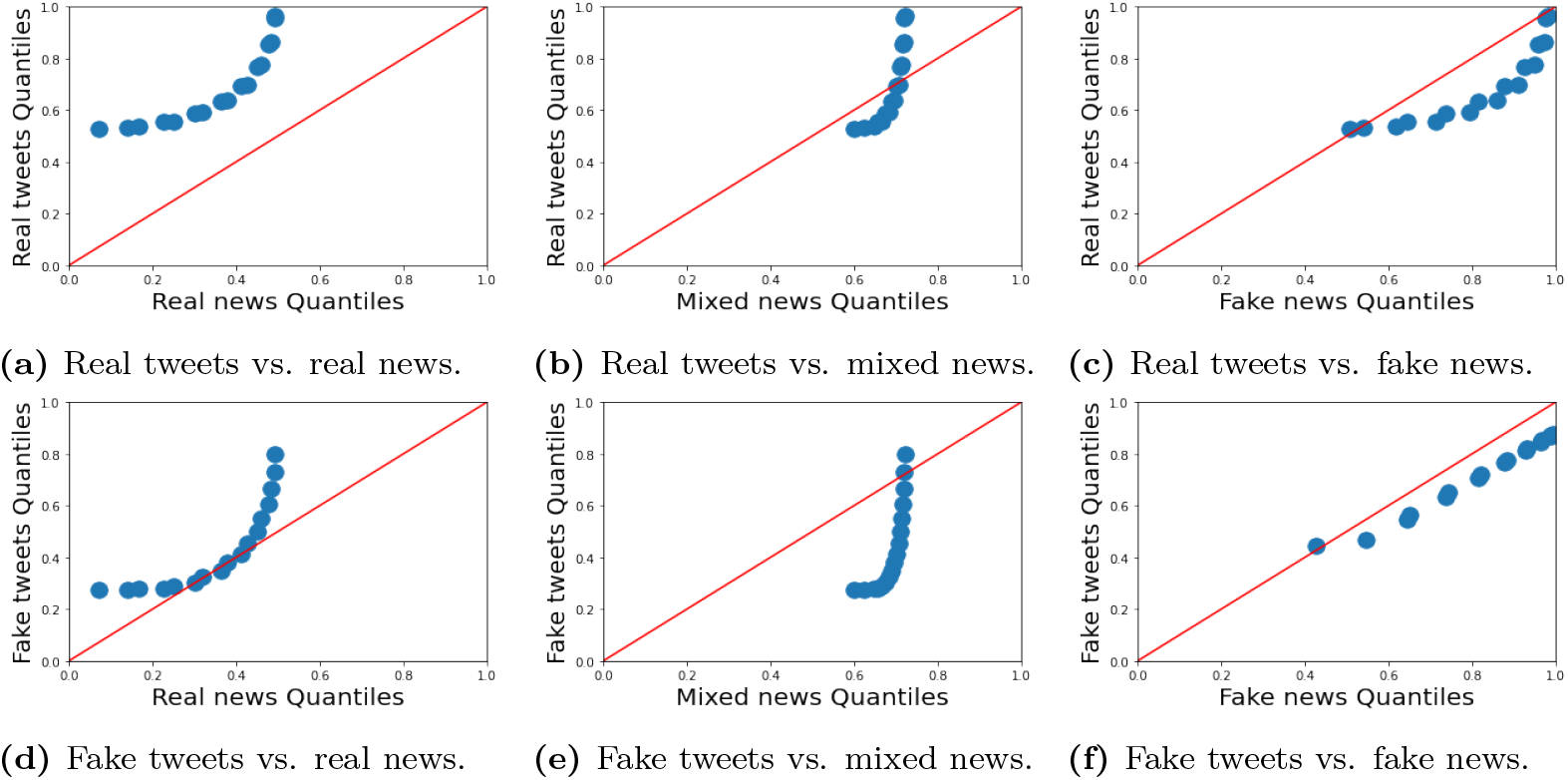
The Quantile-Quantile (Q-Q)-plot of news and tweets used to measure community resilience where x-axis refers to the quantiles of real, mixed, or fake news and y-axis indicates the quantiles of real, fake, or all tweets.

### Summary of Resilience-Related Analysis

We summarize the findings obtained from the discussion above as follows:

- Based on fake news, the public may believe that the community is resilient, which is not the case. Additionally, the results indicate that fake news shares the same viewpoint. They underestimate COVID-19’s adverse effects and demonstrate a higher level of resilience than that measured by real news. This perspective prolongs the time required for actual complete recovery. Further, based on this finding, we observe that fake news is not always pessimistic or negative.
- From community resilience point of view, mixed news is more optimistic than real news showing higher resilience. This may be because mixed news contains fake news, which underestimates the impact of COVID-19.
- Compared to propagated fake tweets, propagated fake news is more unrealistic from community resilience point of view. They demonstrate a greater capacity for community resilience. This finding is reasonable because the source of fake news frequently intends to cause harm, whereas fake news may be spread by people who may have no bad intent but mistakenly believe it or have no knowledge to judge the information credibility.
- From community resilience point of view, propagated real news is slightly more negative than original real tweets, showing a lower level of community resilience. This is because the original intent of fake news originators has been diluted through the process of propagation.

## Conclusion

This section summarizes the key contributions made in this work and answers the research questions raised in Section “Research Goal, Contributions, and Questions”. In addition, we suggest future research directions.

### Summary of the Key Contributions

In this paper, we analyzed community resilience (CR) during the COVID-19 pandemic in the US from Feb. 2020 to Jun. 2021 based on both news articles and tweets on social media. We measured CR based on two main dimensions developed in this paper: community wellbeing (CW) and resource distribution (RD). We also developed two different dimensions to measure RD: economic resilience and community capital. We leveraged the information provided by fact-checking organizations such as Politifact, Poynter, Snopes, and Factcheck to collect 4,952 full-text news articles and categorize them as real, mixed, or fake news. On the other hand, to identify real and fake tweets, we used the top three machine learning (ML) algorithms among eight ML algorithms being evaluated, i.e., Passive-Aggressive Classifier, Decision Tree Classifier, and AdaBoost Classifier. The three ML algorithms showed at least 95% accuracy in classifying 42,877,312 tweet IDs from Jan. 2020 to Jun. 2021 into fake and real tweets based on the majority rule as our experimental tweets dataset. To improve the operationalization and sociological significance of this work, we used dimension reduction techniques, including linear transformations, nonlinear transformations, and manifold learning to integrate various dimensions of community resilience. We provided the output-oriented and capacity-based resilience analyses for various types of news and tweets and investigated their general trends and relationships. In addition, we evaluated community resilience in terms of the meantime to absorption, community non-functioning, and recovery under various critical community functionality thresholds that determine the deadlock of community failure.

### Answers to the Research Questions

**RQ1**. *What are the main trends observed in community resilience and its key attributes, i*.*e*., *community wellbeing and resource distribution?*

**Answer**. Among the PCAs with various kernel types, the SVD, the isomap, and the Locally Linear Embedding, we used the incremental PCA to integrate dimensions of resource distribution and community resilience due to the higher level of variance information ratio and the preservation of temporal dependency information. In September 2020, CW reached its peak in fake tweets and real/fake news. The peaks of CW in real tweets and mixed news, on the other hand, occur in February 2020 and June 2021, respectively. Additionally, we observe that CW reaches a low point by the end of 2020 when real tweets are used. Plus, the findings suggest that the resource distribution trends observed in mixed/fake news and real/fake tweets are comparable to those observed in community capital. On the other hand, the trend in real news about resource distribution corresponds to the trend in economic functionality. Take note that both real and fake news follow the same pattern in terms of resource distribution and community wellbeing. Community resilience trends in real/fake news and real/fake tweets are comparable to resource distribution trends. On the other hand, trends in real, mixed, and fake news regarding resource distribution are similar to the trends in community wellbeing. Fake news has a more even distribution of resources than real news. Finally, fake news has a higher community resilience than real news. By creating unrealistic optimism about the future, fake news has the potential to mislead people into taking inappropriate actions in response to the COVID-19.

**RQ2**. *What are the key differences and correlations between the community resilience measured on various types of news and tweets?*

**Answer**. According to the findings, fake tweet articles have an 80% probability of correlating positively with fake news for resilience-related characteristics. Additionally, Pearson and Kendall tau correlations indicate that the correlation between fake news and real tweets is negative, with an 80 percent probability. Additionally, we discovered that mixed news has a 95% probability of negatively correlating with real and fake tweets across all types of CR attributes. Statistical hypothesis tests, both parametric and non-parametric, demonstrate the distribution’s similarity across multiple scenarios. We observe that fake and real tweets have a 60% probability of having similar distributions. This implies that fake tweets can accurately reflect the actual state of community resilience (CR), regardless of their veracity.

**RQ3**. *What are the level of the community resilience metrics, e*.*g*., *absorption and recovery during COVID-19 on various types of news and tweets?*

**Answer**. According to Tables 6-10, both fake and mixed news exhibit a greater level of absorption than real news for all critical CF threshold values and resilience-related characteristics. Fake news has the highest level of absorption for all critical threshold values, both in terms of community well-being and economic functionality. The number of months that the community is unable to function (TNF) varies between 0 and 17 months, depending on the critical threshold value. For real news, TNF is equal to 17 months when *b* = 0.5. Fake news typically exhibits the greatest degree of recovery in terms of community functionality, economics, community capital, and resource distribution. As a result, we can conclude that mixed/fake news frequently underestimates COVID-19’s negative impact on the community. The negative consequences of fake news complicate not only the handling of COVID-19, but also the recovery process.

### Future Research Directions

We suggest the following future research directions.

First, in this work, we only used Twitter to gather all real and fake news to investigate the behavior of the population. One should be extremely careful in analyzing social media information. Surveys can provide high-quality data for analyzing population behavior, albeit at a cost. Additional research can be conducted to examine the correlations between fake/real news and survey responses. Nonetheless, considering additional social media platforms for future research may be beneficial.

Second, while we propose in this work to quantify community resilience using social media data (e.g., tweets), we are still in the first phase of this journey, namely enhancing community resilience. The literature does not include a thorough examination of the prediction models for community resilience. As a result, more sophisticated models are required to forecast how distinct communities will respond to a variety of events and epidemics. It specifically calls for developing a multi-agent model that accounts for the spread of fake news. The approach described in this work must be extended further to validate the model. The next step on this path is to predict output-oriented community resilience using machine and deep learning techniques.

Third, we consider community capital and economic resilience as resource distribution metrics in this work. Apart from these metrics, institutional and infrastructure resilience are also critical aspects of resource distribution that can have an effect on community resilience. Additionally, we use anxiety, anger, and sadness to ascertain the community’s level of wellbeing. Additional metrics for wellbeing can be added. This necessitates the development of new techniques for assessing additional potential indicators of community wellbeing.

Finally, one can choose appropriate engagement strategies, such as collaborative adaptive management and joint fact-finding, based on a community’s social characteristics and the perspectives of its stakeholders, in order to determine appropriate policies to enhance community resilience.

## Data Availability

https://github.com/Jab-V/Community-Resilience-to-COVID-19-Using-Social-Media

